# Inequalities in oral health: Estimating the longitudinal economic burden of dental caries by deprivation status in six countries

**DOI:** 10.1101/2024.02.12.24302677

**Authors:** Gerard Dunleavy, Neeladri Verma, Radha Raghupathy, Shivangi Jain, Joao Hofmeister, Rob Cook, Marko Vujicic, Moritz Kebschull, Iain Chapple, Nicola West, Nigel Pitts

**Affiliations:** Economist Impact, UK; Division of Haematology, Medical Oncology & Haematopoietic Stem Cell Transplantation, Department of Medicine, The University of Hong Kong, Hong Kong; Health Policy Institute, American Dental Association; Birmingham NIHR Biomedical Research Centre, and Institute of Clinical Sciences College of Medical & Dental Sciences, The University of Birmingham, UK; Division of Periodontics, Section of Oral, Diagnostic and Rehabilitation Sciences, College of Dental Medicine, Columbia University, New York, NY, USA; Bristol Dental School, University of Bristol, UK; Faculty of Dentistry, Oral & Craniofacial Sciences, King’s College London, UK

**Author notes:** contributed equally (Nicola West, Nigel Pitts).

**Keywords:** Oral health, dental caries, inequalities, economic burden, prevention, non-communicable diseases, public health, socioeconomic

## Abstract

**Background:** The recent World Health Organization (WHO) resolution on oral health urges pivoting to a preventive approach and integration of oral health into the non-communicable diseases agenda. This study aimed to: 1) explore the healthcare costs of managing dental caries between the ages of 12 and 65 years across socioeconomic groups in six countries (Brazil, France, Germany, Indonesia, Italy, UK), and 2) estimate the potential reduction in direct costs from non-targeted and targeted oral health-promoting interventions.

**Methods:** A cohort simulation model was developed to estimate direct costs of over time for different socioeconomic groups. National-level DMFT (dentine threshold) data, the relative likelihood of receiving an intervention (such as a restorative procedure, tooth extraction and replacement), and clinically-guided assumptions were used to populate the model. A hypothetical group of upstream and downstream preventive interventions were applied either uniformly across all deprivation groups to reduce caries progression rates by 30% or in a levelled-up fashion with the greatest gains seen in the most deprived group.

**Results:** The population level direct costs of caries from 12 to 65 years of age varied between US10.2bn in Italy to US$36.2bn in Brazil. The highest per-person costs were in the UK at US$22,910 and the lowest in Indonesia at US$7,414. The per-person direct costs were highest in the most deprived group across Brazil, France, Italy and the UK. With the uniform application of preventive measures across all deprivation groups, the greatest reduction in per-person costs for caries management was seen in the most deprived group across all countries except Indonesia. With a levelling-up approach, cost reductions in the most deprived group ranged from US$3,948 in Indonesia to US$17,728 in the UK.

**Conclusion:** Our exploratory analysis shows the disproportionate economic burden of caries in the most deprived groups and highlights the significant opportunity to reduce direct costs via levelling-up preventive measures. The healthcare burden stems from a higher baseline caries experience and greater annual progression rates in the most deprived. Therefore, preventive measures should be primarily aimed at reducing early childhood caries, but also applied across all ages.

## Background

Dental caries affect about 2 billion people worldwide and is the most common non-communicable disease.^1, 2^ The main risk factors for caries include a high-sugar diet, poor oral hygiene, and inadequate exposure to fluoride — all of which are preventable.^3^ Yet, use of preventive approaches to care are lacking and the burden of caries is rising. The total number of individuals with caries at the untreated dentine cavity threshold has risen by 46% between 1990 and 2019; mostly attributable to population change, urbanisation, and lifestyle changes.^4^

The impact of the rising caries prevalence is disproportionately higher among socioeconomically deprived groups. The Global Burden of Disease (GBD) 2019 data showed that caries prevalence in both deciduous and permanent teeth was higher in countries with a lower Social Development Index (SDI).^2^ A systematic review of 41 studies, including adults between 19 and 60 years of age, showed that lower educational attainment, income, socioeconomic status at an individual level, and a higher Gini coefficient at the country level were associated with a higher risk of caries.^5^ Whilst gaps exist in our understanding of the drivers of these inequalities, at an individual level, lifestyle factors, lack of awareness, inadequate oral hygiene, and poor access to care are major drivers.

Additional systematic societal factors may also drive inequalities. For example, people in deprived areas are more likely to consume foods and beverages with higher refined sugar content.^6^ The marketing activities of private companies promoting tobacco, alcohol, high-sugar sweets and beverages more purposefully target lower-middle income countries, emerging economies and deprived populations.^1^ These population segments are particularly vulnerable due to food insecurity, poor access to nutritious food choices, and may also be dependent on these companies for employment and income.^7^ Foods with a high refined sugar content are more readily available in deprived neighbourhoods and usually cheaper than more nutritious alternatives. Thus, the cycle of malnutrition and caries continues in the 21^st^ century.^8^ Deprived populations also have poor access to oral health promotion efforts and caries care.^9^

Both upstream and downstream preventive measures have been shown to lower caries risk. Community-level water fluoridation is a cost-effective population health strategy to reduce caries.^10, 11^ Data from a cross-sectional study in the United States has shown that water fluoridation can also narrow the gap in caries prevalence between different socioeconomic classes in children with deciduous teeth.^12^ However, children in lower socioeconomic groups are less likely to live in localities with fluoridated water.^12^ The institution of extra taxation on high-sugar drinks has also demonstrated efficacy in lowering sugar consumption and caries risk at a population level, which could potentially be more impactful among the most socioeconomically deprived in society.^13, 14^ However, only 57% of the global population lives in countries where sugar tax is implemented. Furthermore, the applied excise tax is relatively small, accounting for approximately 6.6% of the price of the most-sold brand of carbonated drink. Additionally, 46% of countries that impose taxes on high-sugar drinks also apply similar taxation to unsweetened bottled water.^1^

At an individual level, improved oral hygiene and reduced consumption of foods and drinks that contain high-sugar levels are key factors in reducing caries^15^. Tooth brushing twice daily with toothpaste containing 1000-1500ppm of fluoride, albeit a simple measure, remains inaccessible to many low-income communities.^16^ The application of topical fluoride varnish forms a protective layer of fluorapatite on teeth, which can resist demineralisation and further enhance remineralisation, preventing caries progression.^17^ Fissure sealants work as a physical barrier to seal deep fissures preventing food and bacterial accumulation with subsequent caries formation.^17^ A study performed in the North West of England between 1990 and 1999 showed that children from more deprived communities were less likely to receive professional application of fluoride varnish; yet, there is a lack of more recent data on this important preventive measure.^18^ While preventive measures are effective in lowering the caries burden, they remain less accessible to the most deprived, who remain the most severely impacted.

There is limited data on the economic burden of caries and the available information is dated. Global estimates in 2015 suggested that the total worldwide costs of oral and dental diseases amounted to approximately US$544bn. Of this, approximately 45% ($245bn), was estimated to be due to caries.^19, 20^ Data from England in 2015-2016 showed that £50.5m was spent on children less than 19 years of age for tooth extractions, a majority of which were due to tooth decay.^21^ Furthermore, there is no data on the long-term costs of caries of permanent teeth occurring in children. The effect of instituting early prevention and management measures on bridging socioeconomic inequities and reducing direct costs remains to be studied.

Here we report a Caries Prevention and Care Cost Calculator that we have developed to: 1) longitudinally determine the direct costs of management of dental caries between 12 and 65 years of age and 2) the potential reduction in direct costs from non-targeted and targeted oral health-promotion interventions across different socioeconomic groups in six countries (Brazil, France, Germany, Indonesia, Italy, and the United Kingdom (UK). The countries were chosen to generate a representative sample across parameters such as per capita income, population size, levels of inequality, caries prevalence, and structural features of health systems.

## Methods

A simplified conceptual framework was developed to demonstrate the progression of caries from a healthy tooth to an unsalvageable carious tooth that requires extraction. The stages in the progression include the development of an initial white spot lesion caused by demineralisation followed by established tooth decay (caries). If left untreated, caries can progress to involve the dental pulp (root canal system) and the tooth may eventually become unsalvageable (Figure 1). The framework also includes interventions for primary prevention at various stages to prevent the development of or limit the progression of caries (secondary prevention). For healthy teeth or those with carious white spot lesions, maintaining good oral hygiene by brushing with fluoridated toothpaste and applying topical fluoride varnish or consuming fluoridated products (e.g fluoridated water or fluoridated salt) are effective preventive measures. Dental fissure sealants effectively prevent caries in healthy teeth or halt the progression of initial carious lesions by sealing the deep grooves and fissures on chewing surfaces, which are prone to decay due to microbial plaque and food accumulation. After established decay has set in and formed a cavity, the caries process cannot be reversed, and management moves to a “restorative/reparative cycle”. Initially, the tooth can be restored using fillings. If the decay progresses to compromise the dental pulp, the tooth will require root canal treatment and/or a crown. If left untreated, the decay may result in an unsalvageable tooth, necessitating extraction followed by replacement, frequently with a dental implant, if affordable, feasible and clinically appropriate.

**Figure 1:**
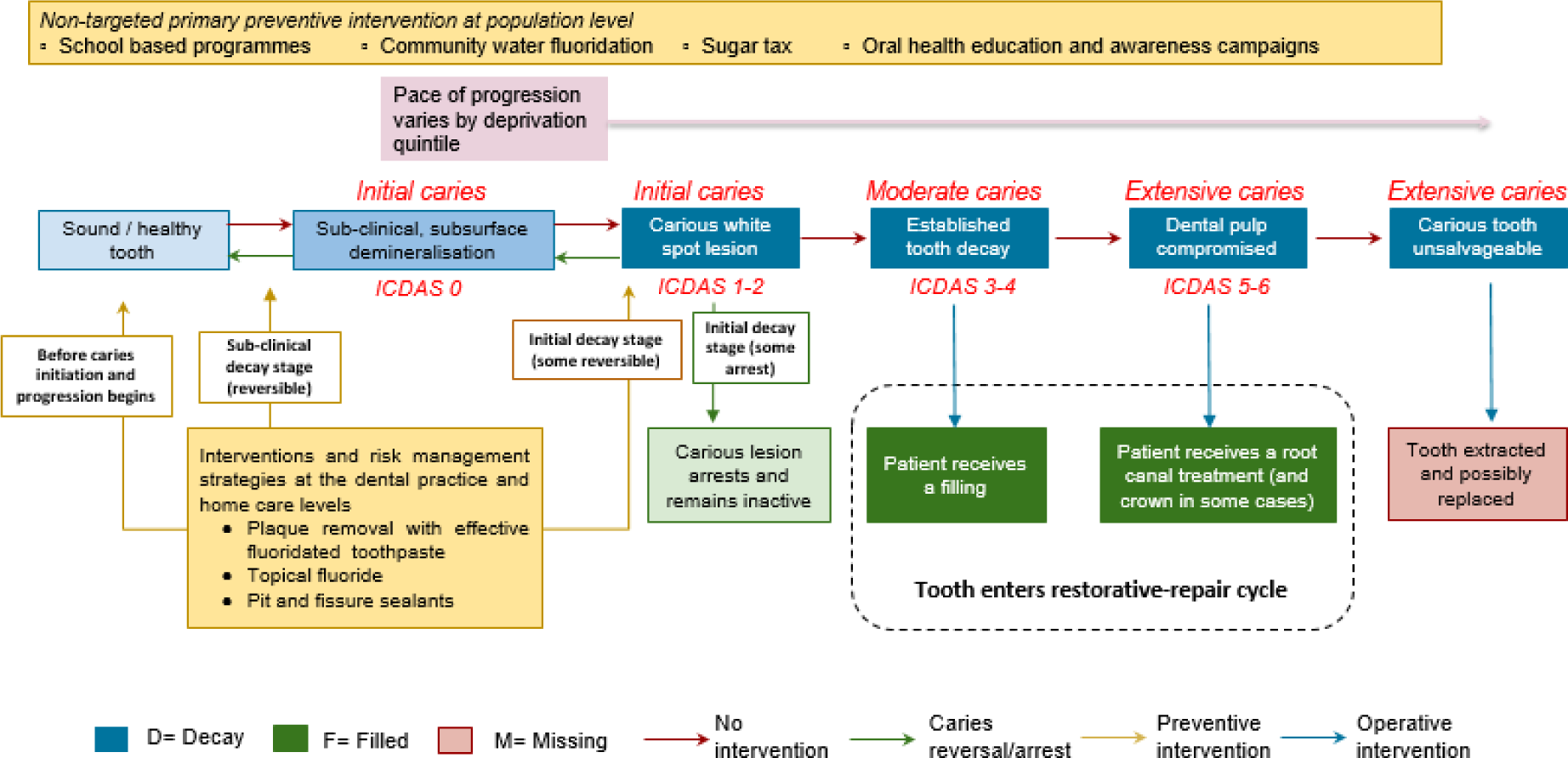
Conceptual framework of dental caries pathway of care

Based on this conceptual framework, we developed a simplified approach to the dental caries clinical care pathway, based on available data, to enable an estimation of the direct costs of dental caries (Figure 2).^22–24^

**Figure 2:**
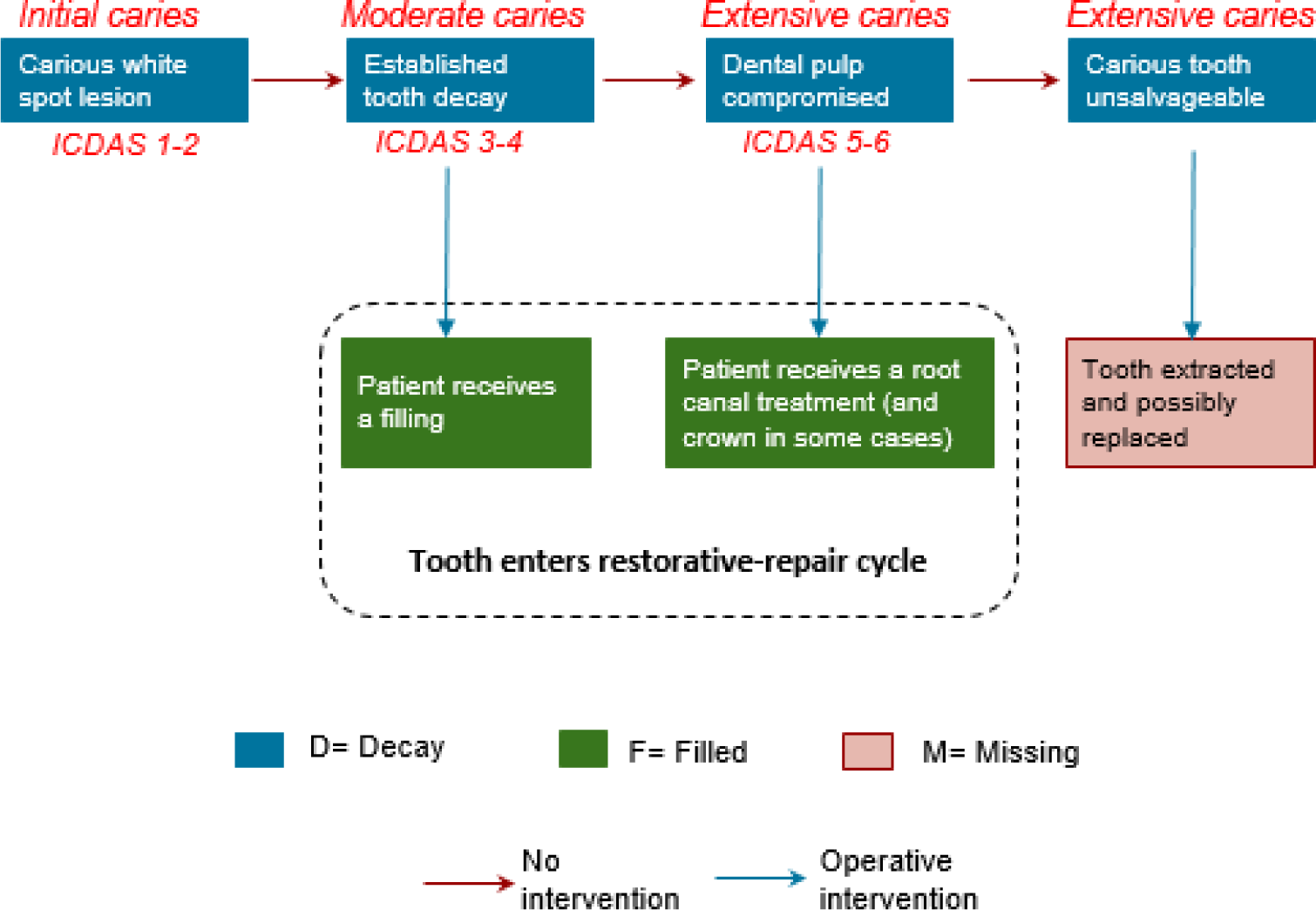
Framework for analysis

### Data inputs Population data

Data from the World Population Prospects (WPP, 27^th^ edition) was used to determine the population size and age structure for each country, broken down into 5-year age groups.^25^ The latest population size estimates for the 10-14 years age cohort were used as the baseline, and projections were made for the population size as the cohort progressed into the 60–64-year age group, based on general population mortality. The death rate assumptions were derived from the WHO data on probability of dying at a specific age.^26^ The formula applied was as follows:

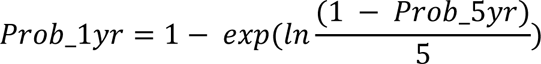

Where:

*Prob_5yr*: The probability of dying between 5-year age cohorts
*Prob_1yr*: The probability of dying at a specific age
*exp*: exponential
*ln*: natural logarithm

### Disaggregation by deprivation quintiles

The English Index of Multiple Deprivation IMD 2019 was used to disaggregate the population in each 5-year age cohort into deprivation quintiles. The quintiles were classified as least deprived, second least deprived, middle deprived, second most deprived and most deprived.^27^ Following the relative ranking system of the IMD 2019, we assigned 20% of the population in each age cohort into each deprivation group.

#### Caries experience data

We used the commonly used population-based measure of DMFT assessed at the caries into the dentine threshold to quantify the current and past caries experience. DMFT refers to the number of decayed, missing and filled permanent teeth and is calculated as the sum of all decayed, missing and filled teeth among individuals in a specific age group divided by the total population in that age group.^28, 29^

DMFT scores for 12-year-olds were sourced from national oral health surveys from Brazil, France, Germany, Indonesia, Italy, and the UK and used as the baseline DMFT/ caries experience (see Table 1). ^30–35^ The UK data is extrapolated from a national survey of England, Wales and Northern Ireland. The total caries experience within the age group was calculated by multiplying the average DMFT scores by the population size in that specific age group.

**Table 1:** Average number of decayed, filled and missing teeth among 12-year-olds by country.

|  | Decayed teeth (DT) | Filled teeth (FT) | Missing teeth (MT) | Decayed, missing or filled teeth (DMFT) |
| --- | --- | --- | --- | --- |
| **Brazil** | 1.12 | 0.73 | 0.12 | 2.07 |
| **France** | 0.5 | 0.6 | 0.13 | 1.23 |
| **Germany** | 0.1 | 0.3 | 0.1 | 0.5 |
| **Indonesia** | 1.8 | 0 | 0.1 | 1.9 |
| **Italy** | 0.71 | 0.36 | 0.02 | 1.09 |
| **UK** | 0.4 | 0.3 | 0.1 | 0.8 |

We used separate Welsh and German data among 12-year-olds that showed a similar 0.4 difference in DMFT scores between the least and most deprived groups when stratified by socioeconomic status.^36, 37^ This difference of 0.4 between deprivation groups was extrapolated to all the studied countries due to the lack of other country specific data.

The average DMFT data reported in each country’s national oral health survey was assumed to apply to the middle-deprived quintile. We derived conversion factors from the Welsh data and adjusted them according to the country’s Gini coefficient relative to the UK, which were used to adjust the DMFT scores for the other quintiles (see Table 2).^38^

**Table 2:** Average DMFT in 12-year-olds by country and income quintile.

|  | Most deprived | Second most deprived | Middle deprived | Second least deprived | Least deprived |
| --- | --- | --- | --- | --- | --- |
|  | Quintile 1 | Quintile 2 | Quintile 3 | Quintile 4 | Quintile 5 |
| **Brazil** | 5.37 | 4.27 | 2.07 | 2.07 | 1.66 |
| **France** | 2.03 | 1.77 | 1.23 | 1.23 | 0.98 |
| **Germany** | 0.78 | 0.69 | 0.50 | 0.50 | 0.40 |
| **Indonesia** | 3.53 | 2.99 | 1.90 | 1.90 | 1.52 |
| **Italy** | 1.88 | 1.62 | 1.09 | 1.09 | 0.87 |
| **UK** | 1.28 | 1.12 | 0.80 | 0.80 | 0.64 |

The calculation used was:

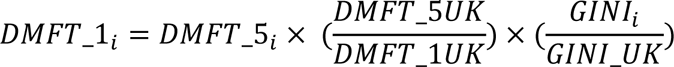

Where:

*DMFT_1_i_* is the DMFT score for the most deprived quintile in country i;
*DMFT_5i_i_* is the DMFT score for the least deprived quintile in country i;
*DMFT_1UK* is the DMFT score for the most deprived quintile in the UK;
*DMFT_5UK* is the DMFT score for the least deprived quintile in the UK;
*GINI_i_* is the Gini coefficient for country i;
*GINI_UK* is the Gini coefficient for the UK

### Caries progression

Data from a large systematic review and meta-analysis reported an unadjusted annual increase in DMFT scores of +0.18 with a lower progression rate of +0.07 after adjusting for preventive interventions.^39^ Based on these data, an annual increment in DMFT score of +0.18 was applied to the middle deprivation cohort and that of +0.07 to the least deprived cohort. We assumed that the least deprived cohort with best access to interventions would have the lowest annual progression rate. Progression rates were then assumed to evolve across income brackets linearly to arrive at the rate of annual progression of DMFT scores:

● Least deprived quintile: +0.07
● Second least deprived quintile: +0.125
● Middle deprived quintile: +0.18
● Second most deprived quintile: +0.235
● Most deprived quintile: +0.29

We assumed that the progression rate in dental caries remains the same across countries and an individual’s lifetime, irrespective of the baseline caries experience.

The overall annual progression rate in DMFT was disaggregated across decayed, filled and missing teeth, based on the progression of decayed, filled and missing teeth values reported in The Dunedin Multidisciplinary Health and Development Study.^40^ We assumed that the distribution of the values reported in the Dunedin study was representative of the middle-deprived quintile. We then adjusted the distribution of individual DMFT values to other quintiles relative to the middle quintile, employing assumptions related to the likelihood of receiving a filling versus extraction across deprivation groups. For instance, individuals in the most deprived group are more likely to receive an extraction rather than preventive management or restorative treatments such as fillings, bridges and implants, even in countries with publicly funded dental care, such as the UK.^41, 42^ The assumptions used in the distribution of DMFT progression between age cohorts across deprivation quintiles are detailed in Table 1 in Additional file 1.

### Direct costs of managing caries

An estimation of the direct costs of managing caries was derived through the triangulation of information gathered from country experts and cost data sourced online. The costs associated with dental caries management per tooth are detailed in Table 2 in Additional file 1. Given the variation in the provision of subsidised care across countries and the lack of information regarding health care system costs in the public sector, private treatment costs in each country were used as a proxy to estimate the direct costs of caries.

An increase in the ‘Fillings’ component of the DMFT by 1 implies that the patient received a new filling. We assumed the need for re-restoration of a filling every 10 years, based on a conservative estimate of the median survival rate of composite fillings.^43, 44^ Of those that received a filling, 9.3% were assumed to have had a root canal treatment, based on data reported in a systematic review.^44^ Among patients receiving a root canal treatment, a proportion were assumed to have also received a crown. Both root canal and crown interventions were weighted such that they were more common among the least deprived, owing to the cost of the procedures. The assumptions applied to the provision of root canals and crown across deprivation groups are detailed in Table 3 in Additional file 1.

An increase in the ‘Missing’ component of DMFT by a value of 1 implies that the patient underwent a single tooth extraction. Following extraction, we assumed that less deprived groups were more likely to receive single tooth implants. Besides dental implants, alternative interventions such as dental bridges or single-tooth removable partial dentures can be used to replace a missing tooth. We included an alternative replacement in the analysis in a treatment-agnostic approach to account for various treatment options that were lower cost to a dental implant, and were more likely to occur in the more deprived groups. To provide a credible range for the estimates and to acknowledge the likelihood that not all patients will receive a replacement for a missing tooth, 60% and 30% of the most and second most deprived groups, respectively, do not receive any replacement for a missing tooth in this analysis. The assumptions applied to the provision of replacements across deprivation groups are detailed in Table 4 in Additional file 1.

Direct costs for caries in each age group were calculated as the cost of treatment per tooth multiplied by the number of teeth requiring the treatment multiplied by the percentage of each deprivation group that are assumed to have the treatment. A 3.5% discount rate for future costs was applied to the calculation based on the UK’s National Institute for Health and Care Excellence (NICE) recommendation.^45^

### Scenario analysis

Two scenario analyses were performed to assess the decrease in per capita costs between 12-65 years of age based on the following interventions:

● Scenario 1 – Application of non-targeted interventions with a decrease in caries progression rates by 30% across each deprivation quintile. The 30% decrease in the caries progression rate was considered conservative, given that the majority of dental caries is preventable via a range of effective public health interventions, such as public water fluoridation, salt fluoridation, reduced sugar consumption (via the implementation of, for example, sugar taxes or enhanced food labelling) and twice daily brushing with fluoridated toothpaste.^46^
● Scenario 2 – A ‘levelling-up’, or proportionate universalism approach, with the scale of prevention and management interventions proportional to the degree of need across deprivation quintiles. In this scenario, the caries progression rate of the least deprived quintile was applied across all quintiles.

## Results

The population-level costs of caries for the current cohort of children aged 12 years projected to 65 years varied between a low of US$ 10.2bn in Italy to a high of US$ 36.2bn in Brazil (Table 3). Of the countries studied, Indonesia is the most populous followed by Brazil. However, in terms of population-level costs, Indonesia ranked second to Brazil with a cost of US$ 26.2bn. The cost of all procedures (except implants and alternative replacements) was estimated to be lower in Indonesia than in Brazil, which may explain the lower direct costs of caries at a population-level in Indonesia, compared to Brazil. The largest per-person costs were estimated in the UK, at US$ 22,910, and the lowest per-person costs in Indonesia, at US$ 7,414.

**Table 3:**
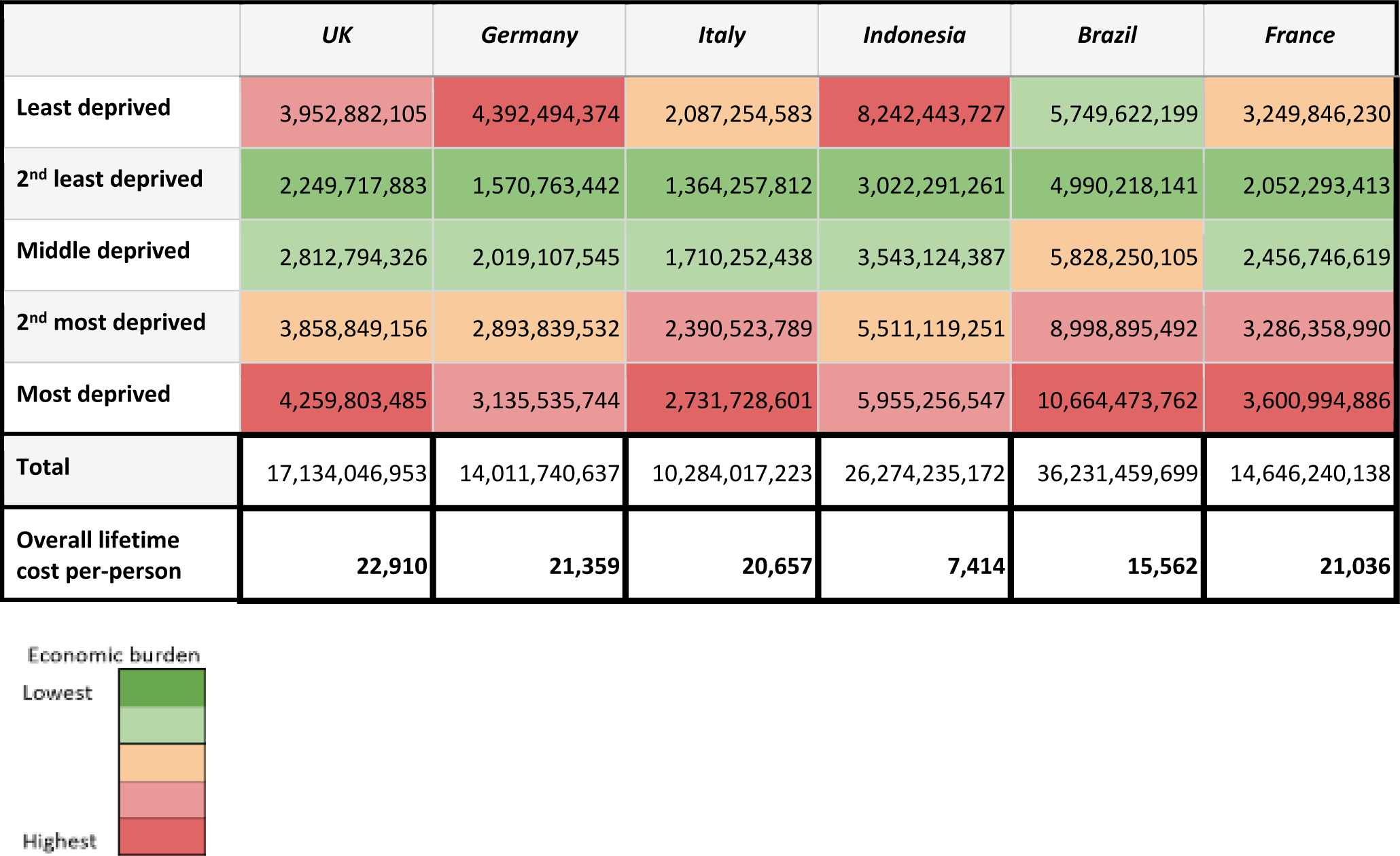
Longitudinal direct costs of caries from 12 to 65 years of age (US$)

|  | *UK* | *Germany* | *Italy* | *Indonesia* | *Brazil* | *France* |
| --- | --- | --- | --- | --- | --- | --- |
| **Least deprived** | 3,952,882,105 | 4,392,494,374 | 2,087,254,583 | 8,242,443,727 | 5,749,622,199 | 3,249,846,230 |
| **2<sup>nd</sup> least deprived** | 2,249,717,883 | 1,570,763,442 | 1,364,257,812 | 3,022,291,261 | 4,990,218,141 | 2,052,293,413 |
| **Middle deprived** | 2,812,794,326 | 2,019,107,545 | 1,710,252,438 | 3,543,124,387 | 5,828,250,105 | 2,456,746,619 |
| **2<sup>nd</sup> most deprived** | 3,858,849,156 | 2,893,839,532 | 2,390,523,789 | 5,511,119,251 | 8,998,895,492 | 3,286,358,990 |
| **Most deprived** | 4,259,803,485 | 3,135,535,744 | 2,731,728,601 | 5,955,256,547 | 10,664,473,762 | 3,600,994,886 |
| **Total** | 17,134,046,953 | 14,011,740,637 | 10,284,017,223 | 26,274,235,172 | 36,231,459,699 | 14,646,240,138 |
| **Overall lifetime cost per-person** | **22,910** | **21,359** | **20,657** | **7,414** | **15,562** | **21,036** |

When disaggregated by deprivation quintile, the most deprived group had the highest per person costs in the UK, Italy, Brazil and France (Table 4). In all countries, the most deprived population had the highest baseline caries experience and the highest rate of progression. While we modelled the most deprived to be less likely to receive expensive treatments, such as root canals and implants and more likely to have extractions, they still had the highest direct costs across four of six countries studied. Of these four countries, Brazil had the greatest difference in costs between the most and the least deprived populations, at US$ 10,555 per person. This can be attributed to Brazil having the higher baseline DMFT value and the greatest inequality among deprivation groups, based on the Gini coefficient. In Germany and Indonesia, the least deprived group had the greatest per-person costs, followed by the most deprived group. Our modelling assumes that the most deprived are more likely to receive just an extraction and a less optimal, lower-cost replacement or no replacement at all, while the least deprived are more likely to get an implant after the extraction. In the UK, Italy, Brazil and France, the cost of an implant is on average 12 to 35 times more expensive than an extraction. However, the ratios are higher in Indonesia and Germany, at 39 and 80 respectively. This may account for the costs in the least deprived being higher than in the most deprived in these two countries. Across all countries, the second least deprived population was associated with the lowest estimated costs. This was largely owing to a lower health burden than the middle and more deprived groups, coupled with the assumption that fewer people in this deprivation group (relative to the least deprived group) would receive a single tooth implant to replace a missing tooth.

**Table 4:** Cost of caries per-person between 12-65 years by deprivation quintile (US$)

|  | UK | Germany | Italy | Indonesia | Brazil | France |
| --- | --- | --- | --- | --- | --- | --- |
| **Least deprived** | 26,427 | 33,479 | 20,963 | 11,629 | 12,348 | 23,339 |
| **2nd least deprived** | 15,040 | 11,972 | 13,701 | 4,264 | 10,717 | 14,738 |
| **Middle deprived** | 18,805 | 15,389 | 17,176 | 4,999 | 12,516 | 17,643 |
| **2nd most deprived** | 25,798 | 22,057 | 24,008 | 7,775 | 19,326 | 23,601 |
| **Most deprived** | 28,479 | 23,899 | 27,435 | 8,402 | 22,903 | 25,860 |

Table 5 compares the direct costs of caries among the most deprived with the average cost of the least and second least deprived groups. Relative to the two least deprived groups, the direct costs range between 5% and 99% higher for the most deprived groups in Germany and Brazil, respectively.

**Table 5:** Difference in per-person cost of caries for people 12-65 years of age between least and most deprived groups.

|  | UK | Germany | Italy | Indonesia | Brazil | France |
| --- | --- | --- | --- | --- | --- | --- |
| Two least deprived groups (averaged) (US\$) | 20,734 | 22,726 | 17,332 | 7,947 | 11,533 | 19,039 |
| Most deprived (US\$) | 28,479 | 23,899 | 27,435 | 8,402 | 22,903 | 25,860 |
| % increase between most deprived and less deprived groups | 37 | 5 | 58 | 6 | 99 | 36 |

We then modelled the impact of preventive interventions on reducing caries-related direct costs. We conducted a scenario where annual progression rates were reduced by 30%, to account for potential upstream and downstream prevention. The conservative estimate of 30% was intervention-agnostic and applied uniformly to all deprivation groups. With the decrease in progression rates, the greatest reduction in per-person costs for caries management was observed in the most deprived group across all countries except Indonesia, where costs decreased by US$ 1,604 and US$ 1,561 in the least and most deprived groups respectively.

A ‘levelling-up’, or proportionate universalism approach, was also applied as a scenario, where preventive and management interventions were proportionate to the degree of need across deprivation quintiles. In this scenario, an annual caries progression rate of +0.07 that originally pertained to the least deprived group was applied across all quintiles. The per-person the per-person reduction in direct costs ranged from US$ 3,948 in Indonesia to US$17,728 in the UK (Figure 3).

**Figure 3.**
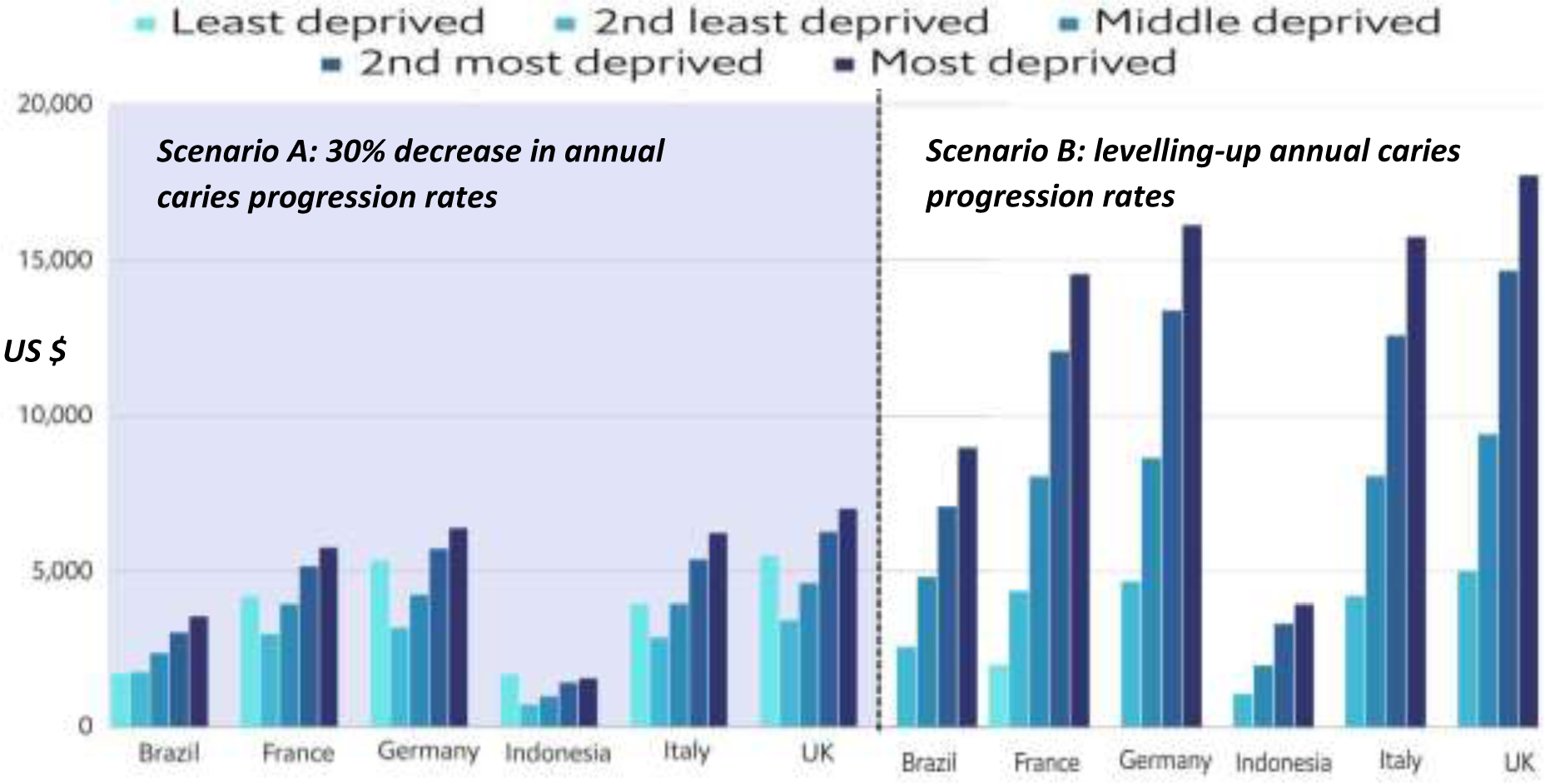
Decrease in per person costs (US$) after uniform application of non-targeted interventions lowering the progression rate by 30% and a levelling up approach to reduce caries progression

## Discussion

To our knowledge, this is the first study to estimate the longitudinal direct costs of caries of permanent teeth occurring in 12-year-olds and how this differs based on socioeconomic status. Despite accounting for the likelihood that more deprived populations receive lower cost and often suboptimal or inappropriate treatment options, such as tooth extraction when a restorative procedure may be more appropriate, the more deprived populations experience a larger long-term economic burden from dental caries. Studies have shown that individuals who are most deprived are more likely to present to the emergency room with non-traumatic dental issues and to be admitted for caries management.^47–49^ Our study has not considered hospitalisation costs; with their inclusion, the difference in direct costs between the most and least deprived is likely to widen. The incorporation of indirect costs, such as transportation to clinics and economic opportunities, would further increase the gap.

The greater caries experience at baseline in the most deprived group is the primary factor leading to an overall higher caries experience and increased direct costs. Therefore, preventive interventions should start early, with a focus on lowering early childhood caries (ECC) and, eventually, caries in young adults and adults.

A multipronged approach, consisting of upstream and downstream efforts, is needed for effective caries prevention. However, there is a dearth of data on the numerical impact of such a holistic approach in lowering the caries burden. The efficacy of various preventive measures has been studied, mostly in isolation. A systematic review of 107 studies showed that community water fluoridation results in a reduction of caries experience by 35% in deciduous teeth and 26% in permanent teeth.^10^ The impact of sugar tax on lowering caries prevalence is less clear, with the majority of evidence derived from modelling studies. A recent umbrella review concluded that a 20% sugar tax would reduce sugar intake by 18% and 20% in low and middle-income and high-income countries, respectively. The review reported that sugar taxes would result in a positive but modest impact on oral health, reducing caries prevalence in children by 2.9% and 2.7% for low and middle-income and high-income countries, respectively, reducing caries counts in adults by 0.03, in both low and middle-income and high-income countries.^50^ The implementation of a sugar tax in the UK has shown a 12.1% relative reduction in hospital admission for carious tooth extractions, but more work needs to be done in estimating the changes in caries prevalence.^51^ There are only a few studies evaluating the impact of school-based oral health education programmes in reducing caries experience. Most failed to show a significant reduction in caries unless combined with fluoride mouth rinses and application of topical fluorides.^52^

At an individual level, tooth brushing with fluoride toothpaste has shown standalone benefit in reducing caries prevalence, while other strategies, such as the application of fluoride varnish, do not appear effective in isolation to reduce caries progression.^53^ A Cochrane systematic review evaluating the impact of tooth brushing showed a 24% reduction in caries experience as measured by DFMS scores among children using fluoridated toothpaste versus those using non-fluoridated toothpaste.^54^

Given the lack of data regarding the decrease in progression rates using a multipronged approach, we assumed a 30% reduction based on diverse literature. With this decrease applied uniformly to all deprivation groups, the most deprived group showed the greatest reduction in costs across all countries, with the exception of Indonesia, where the most deprived group was a close second to the least deprived. A levelling-up approach to prevention resulted in a greater reduction of caries progression in the most deprived group lowered direct costs dramatically for the most deprived.

It should be noted that the most deprived face additional challenges in implementing effective home care strategies to prevent dental caries, especially in times of a cost-of-living crisis. A survey conducted in 2022 by the Oral Health Foundation reported that one in four people in the UK were cutting back on oral health products, such as toothpaste and mouthwash, due to economic challenges.^55^ Even when accessing toothpaste, more deprived populations may not be accessing an effective product. A recent study from Manaus, Brazil, found that cheaper formulations of fluoride toothpaste, which are more commonly used by more deprived populations, lacked a sufficient total fluoride concentration to control dental caries in 92% of the 99 toothpaste tubes tested.^16^

The limitations of our model mainly hinge on challenges in identifying suitable data inputs. national oral health surveys were used to identify the baseline DMFT scores, but these were collected across a range of years (2007-2018). The data does not accurately reflect recent trends and the covid-19 pandemic-related setbacks in oral care. Moreover, the prevalence of caries segregated by deprivation quintile was not available for every country studied.

Annual rates of caries progression across deprivation quintiles have not been documented. Therefore, extrapolations and assumptions had to be made. Studies have shown that the most deprived patients are less likely to get fillings, root canal treatments, or implants but the exact differences in the rates of these treatments between deprivation groups are not known.^41, 42^ Additionally, there is a lack of documentation of health care system costs for caries care in the public sector. Costs from the private sector were used based on triangulation of online data sources and expert input, but there is significant variability in the costs resulting in a wide range of estimates. Our study has used DMFT scores to explore and quantify the health and economic burden of caries, but this score only accounts for established caries and not early caries.

The complete benefit of preventive measures on caries progression can only be estimated by including early, reversible carious lesions. Measures such as the International Caries Detection and Assessment System (ICDAS), the American Dental Association Caries Classification System (ADA CCS), Caries Assessment Spectrum and Treatment (CAST), and Nyvad’s Criteria each facilitate the detection of caries across the entire disease continuum, and their wider use would provide a better estimation of early caries and the cost savings of preventive measures.^56–58^ It is encouraging that the FDI World Dental Federation, having started to promote Minimal Intervention in the Management of Dental Caries in 2002, issued a new Policy Statement in 2019 on Carious Lesions and First Restorative Treatment.^59^ This states that: “FDI World Dental Federation supports a shift in caries management from restorative treatment to measures that arrest and prevent caries development including monitoring, following the concepts of International Caries Classification and Management System (ICCMS)”. Linking of these epidemiological and outcome measures to quality of life metrics would also be of value.

Future studies on the relative costs and benefits of a population approach to oral health and equity should consider collecting the following data for epidemiological and health service research:

● Transparent cost data for publicly funded dental treatment by country (and sub-national or community level)
● Population-level, cross-sectional, data at a country level of the proportion of people paying out-of-pocket for dental care, paying for private treatment through Insurance premiums, and receiving publicly funded dental care through social/worker Insurance or universal coverage. Including mixed schemes or partial charges.
● Longitudinal cohort data on the progression of caries within populations covered by different types of funding schemes.
● New epidemiological and outcome measures for oral health. Metrics that do not weigh a decayed tooth and a filled tooth equally (such as DMFT). Ideally, measures that are:

○ Easy to collect by survey and by health practitioners;
○ Collected (with age-appropriate variations) for all groups across the lifecourse, from preschool children to the elderly;
○ Consider untreated caries, caries experience, treatments received and treatment urgency;
○ Validated against quality of life metrics.

## Conclusion

The cornerstone of dental caries management in dental practice for several decades has been a reparative/restoratively-driven approach that results in significant morbidity and huge costs. This is in stark contrast to preventive models of care that are universally taught at the undergraduate level, and there is increasing emphasis on a more sustainable model of preventive management. Strong data regarding the health and economic benefits of this preventive approach is key to galvanising the support of policymakers.

This exploratory modelling study highlights the impact of caries burden, and how the greatest health burden and direct costs of caries are seen in the most deprived populations. A multipronged preventive approach, if instituted, will offer maximum reductions in direct costs to the most deprived, and highlights a strong case for “levelling-up” preventive actions focused on this segment of the population.

Several data gaps remain to be filled to ensure a more accurate estimation of the value, costs and associated savings of such an approach. Epidemiological measures that include early/reversible caries lesions should continue to be made more user-friendly to facilitate widespread incorporation into such assessments, thereby improving the estimates of benefits offered by preventive care.

The results of this analysis support the case for a more inclusive public health approach to caries management, that incentivises and focuses on prevention and early minimally interventive treatment to improve population-level oral health. Transformative changes in oral healthcare funding models are required to realise the financial benefits of preventive and minimal intervention approaches, and for levelling up oral/dental health to reduce inequalities across socioeconomic groups.

## Contributions

G.D, N.V., and R.R wrote the main manuscript text. G.D, N.V, J.H. prepared the figures. J.H. prepared the tables. N.W. was responsible for the overall strategy, conception and initiation of this research. N.P. outlined the conceptual framework of modern, evidence-based approaches to caries management which was integrated with the new cohort simulation model. I.C. contributed to the overall strategy and concept and critical appraisal of the manuscript. S.J., N.V., G.D. and R.C. developed the analysis framework. G.D, N.V, J.H. collected the data inputs and performed the analysis. All authors read and approved the final manuscript.

## Availability of data and materials

The data related to the assumptions used in the analysis are available in Additional file 1. The dataset generated during this analysis is available from the corresponding author upon reasonable request.

## Abbreviations

ADA CCS: American Dental Association Caries Classification System
CAST: Caries Assessment Spectrum and Treatment
DMFT: Decayed, Missing or Filled Teeth
DT: Decayed teeth
ECC: Early childhood caries
FT: Filled teeth
GBD: The Global Burden of Disease
ICCMS: International Caries Classification and Management System
ICDAS: International Caries Detection and Assessment System
IMD: English Index of Multiple Deprivation
MT: Missing teeth
NICE: National Institute for Health and Care Excellence
SDI: Social Development Index (SDI)
UK: United Kingdom
WHO: World Health Organization
WPP: World Population Prospects

## Additional file 1

**Table 1:** Assumptions used in the distribution of DMFT progression between age cohorts across deprivation quintiles (% of total change in DMFT score attributed to decayed teeth, filled teeth and missing teeth vs middle deprived quintile)

|  | 12 years | 13-18 years | 19-26 years | 27-32 years | 33-38 years | 39-65 years |
| --- | --- | --- | --- | --- | --- | --- |
| ***Least deprived vs middle deprived*** |  |  |  |  |  |  |
| **Decayed** | Baseline | 3.57% | 51.7% | -35% | -43% | -24% |
| **Filled** | Baseline | 96.4% | 42.9% | 110% | 68% | 33% |
| **Missing** | Baseline | 0% | 5.3% | 25% | 75% | 43% |
| ***Second least deprived vs middle deprived*** |  |  |  |  |  |  |
| **Decayed** | Baseline | 5.7% | 56.7% | -37% | 47.5% | 26.3% |
| **Filled** | Baseline | 94.2% | 35.2% | 107% | 62.5% | 27.7% |
| **Missing** | Baseline | 0% | 8.1% | 30% | 85% | 46% |
| ***Middle deprived*** |  |  |  |  |  |  |
| **Decayed** | Baseline | 7.1% | 60.7% | -40% | -50% | 27.7% |
| **Filled** | Baseline | 92.8% | 28.5% | 100% | 50% | 22.2% |
| **Missing** | Baseline | 0% | 10.7% | 40% | 100% | 50% |
| ***Second most deprived vs middle deprived*** |  |  |  |  |  |  |
| **Decayed** | Baseline | 10% | 64.3% | -47.2% | -60% | 30% |
| **Filled** | Baseline | 90% | 21.6% | 92.2% | 45% | 10% |
| **Missing** | Baseline | 0% | 15% | 55% | 115% | 60% |
| ***Most deprived vs middle deprived*** |  |  |  |  |  |  |
| **Decayed** | Baseline | 14.3% | 68% | -55% | -70% | 31.8% |
| **Filled** | Baseline | 85.7% | 12.7% | 75% | 40% | 3.2% |
| **Missing** | Baseline | 0% | 19.3% | 80% | 130% | 65% |

**Table 2:** Costs associated with dental caries management per tooth (2023, in US$, based on currency conversion rates on 12/06/2023)

|  | Filling | Root canal | Crown | Extraction | Implant | Alternative replacement* |
| --- | --- | --- | --- | --- | --- | --- |
| **UK** | 136 | 718 | 669 | 125 | 3133 | 251 |
| **Brazil** | 69 | 213 | 500 | 43 | 540 | 43 |
| **Italy** | 158 | 349 | 460 | 58 | 2,052 | 164 |
| **France** | 105 | 662 | 755 | 93 | 1,980 | 158 |
| **Indonesia** | 25 | 117 | 151 | 26 | 1,010 | 81 |
| **Germany** | 129 | 645 | 1000 | 38 | 3,051 | 244 |

*\*Dental bridges or single-tooth removable partial dentures are alternatives to dental implants in replacing a missing tooth. We included an alternative replacement option in the analysis to account for treatment options that cost lower than a dental implant*

**Table 3:** Provision of root canals and crown across deprivation groups (%)

|  | Root canal | Crown |
| --- | --- | --- |
| Least deprived | 18 | 7.2 |
| 2nd least deprived | 12.5 | 3.75 |
| Middle deprived | 9 | .18 |
| 2nd most deprived | 5.5 | 0 |
| Most deprived | 1.5 | 0 |

**Table 4:** Provision of replacements across deprivation groups (%)

|  | Receive single implant^ | Receive cheaper alternative replacement | No replacement post-tooth extraction* |
| --- | --- | --- | --- |
| Least deprived | 100 | 0 | 0 |
| 2nd least deprived | 80 | 20 | 0 |
| Middle deprived | 10 | 90 | 0 |
| 2nd most deprived | 0 | 70 | 30 |
| Most deprived | 0 | 40 | 60 |

^Assumes patients are suitable candidates for the implants (have good bone support)
\*The cheaper replacement in each country was 8% of the cost of the single implant

